# Implementing a data infrastructure for precision oncology projects leveraging REDCap

**DOI:** 10.1101/2022.05.09.22274599

**Authors:** Charles Vesteghem, Simon Christian Dahl, Rasmus Froberg Brøndum, Mads Sønderkær, Julie Støve Bødker, Alexander Schmitz, Joachim Weischenfeldt, Inge Søkilde Pedersen, Mia Sommer, Anne Stoffersen Rytter, Marlene Maria Nielsen, Morten Ladekarl, Marianne Tang Severinsen, Karen Dybkær, Kirsten Grønbæk, Tarec El-Galaly, Anne Stidsholt Roug, Martin Bøgsted

## Abstract

**Objectives:** To facilitate clinical implementation and research in precision oncology, notably the pairing of patients, variants and treatments to identify candidates for clinical trials, we have built a data infrastructure to 1) capture and store data, 2) reduce manual tasks for clinical and genomic data collection and management, 3) combine data for quality controls, reporting and findability.

**Infrastructure:** The infrastructure uses REDCap repositories to capture and store data. The structure of these repositories is customized for each project. Additionally, a cross-project web platform was developed using software development best practices and state-of-the-art web technologies to circumvent REDCap’s limitations and integrate other third-party resources. Using REDCap’s application programming interfaces, this platform allowed validation of data across multiple repositories, easy import of data from external sources, generation of overviews of included patients and available data, combination of genomic and clinical data to generate tumour board reports and the findability of data. Its design was driven by data stewardship best practices.

**Usage:** Across four precision medicine projects, the infrastructure has been used to collect data for 1921 patients, including 453 genomic data files. The custom-built web platform made it possible to import, validate, and present data in a comprehensive manner. This included building tumour board reports for clinicians, combining clinical and genomic data, and search functionalities for researchers.

**Discussion:** REDCap allowed us to capitalize on the numerous data capture and management features developed in this solution. Designing a cross-project platform guarantees long-term relevance where developments can be mutualised across projects and allowed us to make the overall solution more compliant with the FAIR (Findable, Accessible, Interoperable, Reusable) data principles. Further developments should be considered, notably automatic retrieval of data from electronic health records to limit the number of manual tasks.

**Conclusion:** The proposed infrastructure allowed our precision oncology projects to gain efficiency in data collection and increase data quality by reducing manual work, and it gave a straightforward and customized access to data for researchers and clinicians.

## 1. INTRODUCTION

Due to an improved and ever-growing understanding of the complexity and heterogeneity of cancer diseases, treatment planning and research will rely on an increasing amount of patient and tumour specific data. A streamlined workflow to produce, collect, and report these data will make them easily available and more readily clinically applicable and thereby improve the basis for the planning of treatment at a patient-specific level and facilitate research projects.

In precision oncology, treatment decisions are based on extensive clinical data and detailed genomic profiles of the tumour^1^. Managing and combining these data can be cumbersome and time consuming due to the volume of data and the variety of electronic health record (EHR) systems^2^, ranging from pathology systems to patient journals. The design of treatments for cancer patients requires collaboration of a multidisciplinary team that needs easy access to the most relevant information to arrive at a consensus on the treatment plan. Another crucial aspect to support cancer research is to make the data collected easily searchable, both locally and internationally, as defined in the FAIR (Findable, Accessible, Interoperable, Reusable) data principles^3^. Solutions exist, open source or commercial, that fulfil parts of these needs, notably for genomic data interpretation^4,5^, pathology reporting^6^, or tumour board reporting^7^. However, they lack the flexibility provided by data capture solutions such as REDCap^8^ in terms of data structure, to fit the specific needs of each project. In return, a solution purely based on REDCap repositories will lack necessary features for a fully integrated, end-to-end solution.

We propose here an infrastructure based on REDCap complemented by a modular web platform, referred to in the following as PrOnco (Precision Oncology Platform), intended to: 1) facilitate the capture, storage, and accessibility of relevant information, 2) minimise manual collection and management of clinical and genomic data to reduce workload and improve quality, 3) combine data for quality controls, reporting, and findability.

## 2. INFRASTRUCTURE

This approach was used for four onco-haematology prospective clinical protocols, ProSeq^9^, RetroSeq, ProSeq Cancer (the North Denmark Region Committee on Health Research Ethics approval no. N-1706295, N-20160089, and N-20200018, respectively), and the Danish Research Center for Precision Medicine in Blood Cancers (DCCC-Haem) (Danish National Committee on Health Ethic approval no. 1705391). As an illustration of the architecture put in place, we will be focusing on the ProSeq project in the following.

### 2.1. Inclusions and clinical data

The ProSeq project was conducted at the Department of Haematology, Aalborg University Hospital, where patients with relapse or progression of a haematological neoplasm were offered DNA and RNA sequencing of their tumours to investigate the potential for implementing precision oncology. For this project, the inclusion data and clinical data were stored separately using REDCap v10.6.26 (see Figure 2).

**Figure 1.**
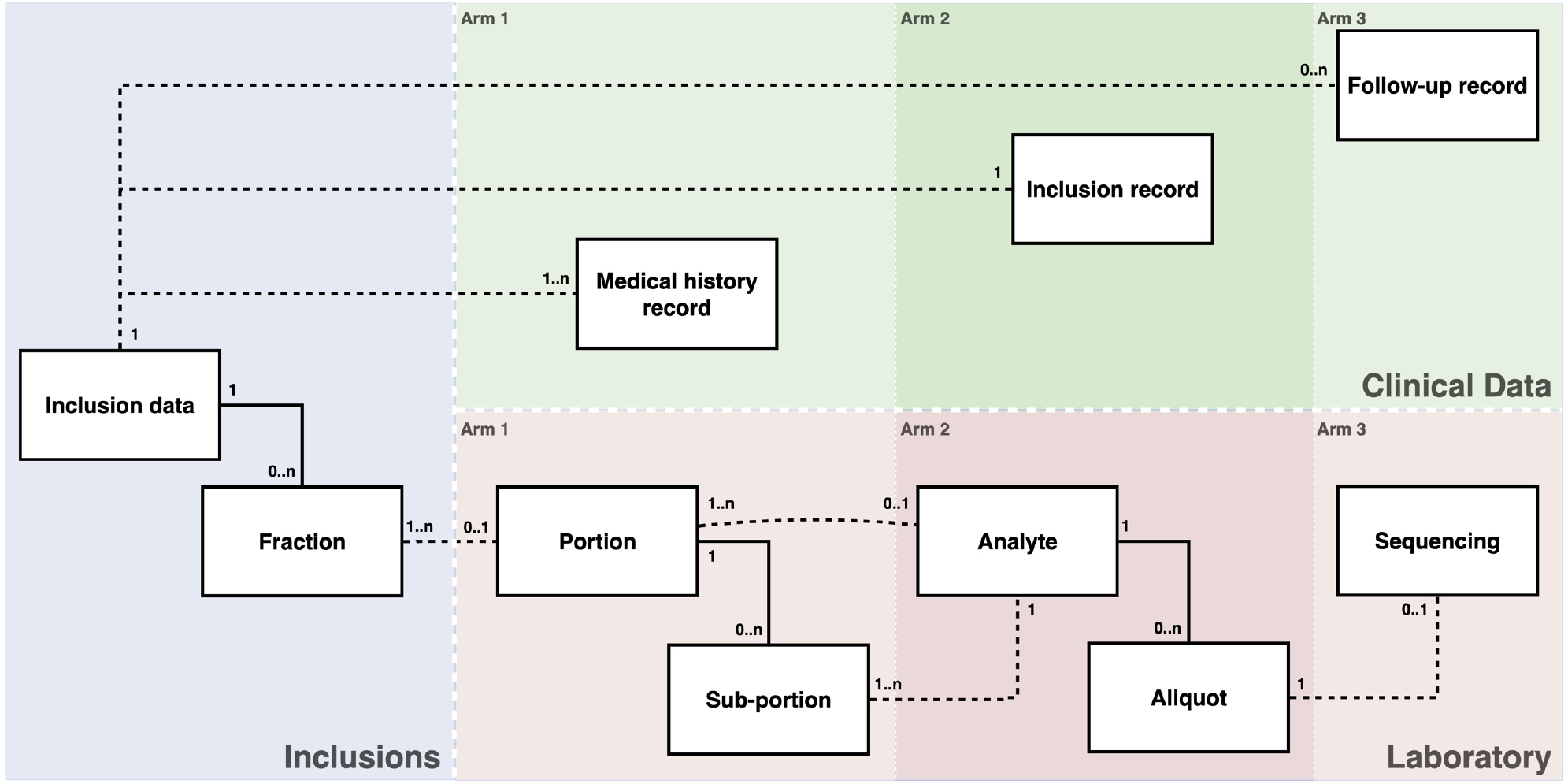
Data structure in REDCap. The three coloured rectangles represent the three REDCap repositories used for the project, the Inclusions, Clinical Data and Laboratory repositories. Within a repository, the records could be subsequently divided into arms as represented with shades of the corresponding colours for the Clinical Data and Laboratory repositories. Solid lines represent structural relations, while dashed lines represent relation using IDs as text.

**Figure 2.**
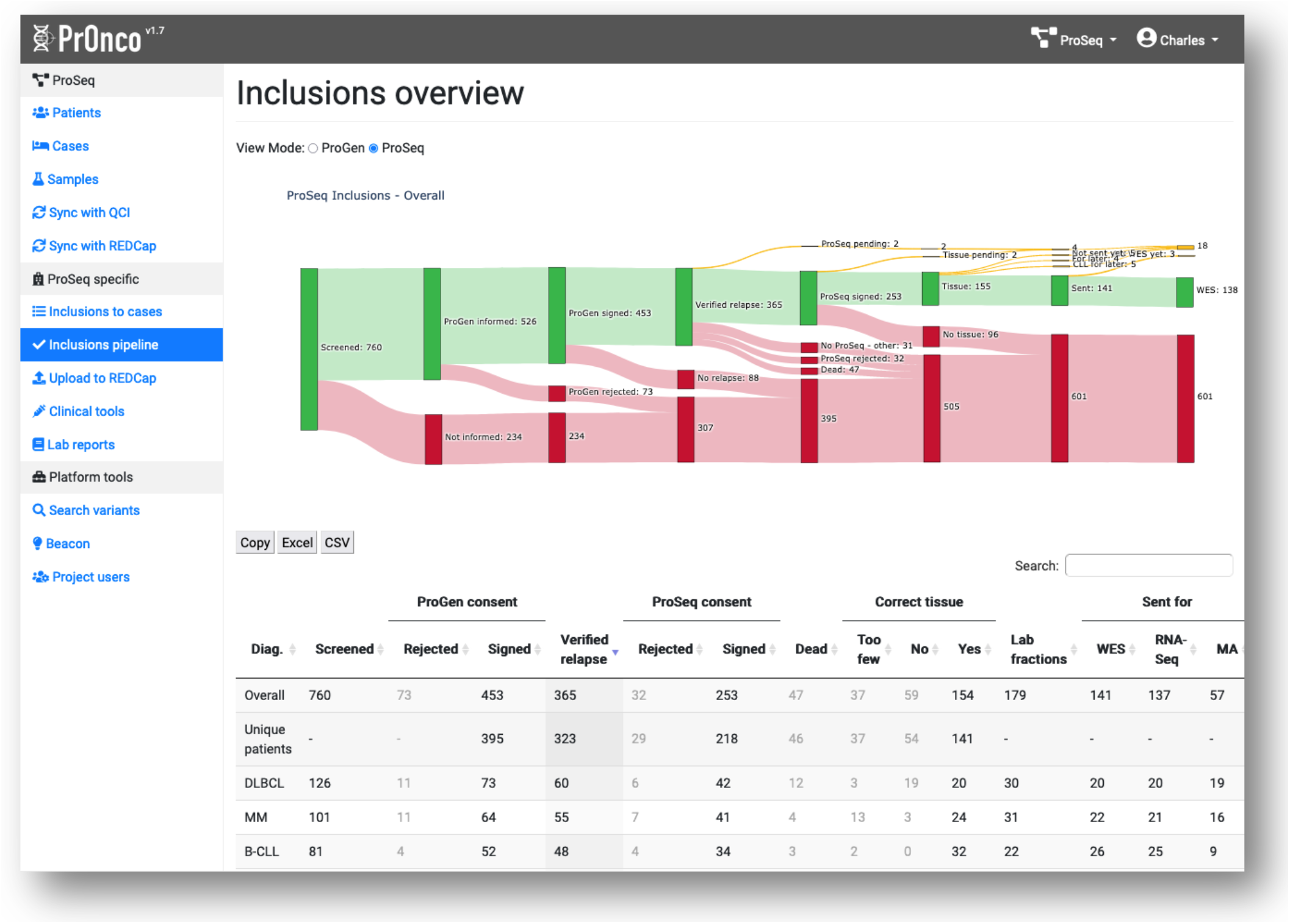
Screenshot of the Inclusions overview. A Sankey chart is used to visualize the drop-out of patients during inclusion and laboratory processing. A listing for each diagnosis type is also provided. The left side bar contains the accessible interfaces available within the selected project. Some of these interfaces are available to all projects, while others are project specific. The top bar displays the logged-in user and the selected project.

The clinical data part consisted of extensive information on the patient trajectory. Individual REDCap records were created for each relapse events where consent was collected. These records are referred to as inclusion records. Additionally, records for all previous events including primary diagnosis and for later relapses where consent could not be collected were created and referred to as medical history records and follow-up records respectively. Each of these records was hereafter referred generically to as clinical records.

To follow the FAIR data principles, variables were, whenever possible, coded according to international standards, such as the International Classification of Diseases, tenth revision (ICD-10)^10^ codes for diagnosis; the International Classification of Diseases for Oncology, third revision (ICD-O-3)^11^ codes for pathology results; and the Anatomical Therapeutic Chemical (ATC) classification ^12^ codes for drugs^13^. Project nurses performed the collection of these data by reviewing patient records manually.

In contrast, for data available in a structured form, i.e., pharmacy data, transfusion data, and outpatient visit data, a set of import tools were built alongside REDCap into the PrOnco web platform to reduce workload. In addition to the import tools, reporting, data synchronization, and validation tools were developed. The purpose of the reporting tool was to give project members a real-time overview of the inclusion process (see Figure 2).

A data synchronization functionality was developed to automatically retrieve and combine the data available in REDCap, in order to store limited information about patients and clinical records in PrOnco, and to enable the storage of annotated genomic data, as described below.

A data validation tool was coupled with the data synchronization functionality to allow project nurses to locate and correct potential inconsistencies in the REDCap data.

### 2.2. Laboratory metadata and genomic data

Information about available tissue for sequencing was stored in the Inclusion REDCap repository. To make the work of the biobank laboratory technicians more efficient, a tool was integrated on the platform to upload this information for included patients directly from an extract from the central Danish pathology biobank: Bio and Genome Bank Denmark (RBGB)^14^.

To record every step of tissue processing, an additional REDCap repository was created to build the backbone of a laboratory information management system (LIMS). The corresponding data were structured following the Genomics Data Commons (GDC)^15^ laboratory metadata structure.

The output from the sequencing was processed in a bioinformatics workflow^9^. Once completed, an extensive number of quality metrics were pushed to a sequencing record in REDCap within the same repository alongside the Variant Call Format (VCF)^16^ file containing the somatic variants. In the ProSeq project, molecular biologists interpreted the clinical relevance of the detected variants using the decision-support tool Qiagen Clinical Insight Interpret (QCI-I)^4^. From this platform, a genomic report was retrieved for each patient. Custom scripts have been developed and implemented to automate this process through an application programming interface (API) (see Figure 3).

**Figure 3.**
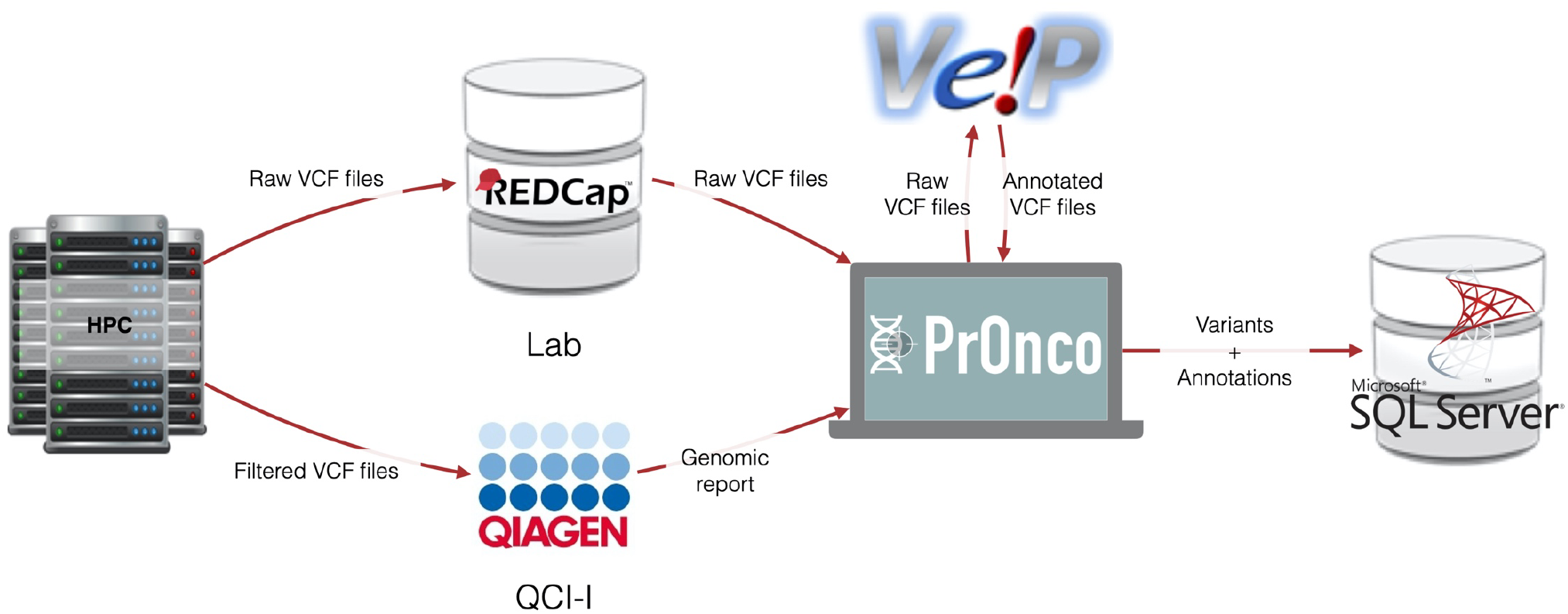
Variant data workflow. This figure illustrates the flow of data once the variants have been called through the bioinformatics pipeline at the Computerome 2.0 high-performance computing centre. All data transfers and data processing steps are automated in this case. A genomic report contains interpretations of the clinical relevance of variants.

Reporting and validation tools have been developed to ensure the quality of the process. The laboratory data validation generates warnings that were reported together with the clinical data issues.

It validates the traceability of tissues and if the necessary data are reported and the identification numbers follow a predefined format.

The main laboratory reporting tool allows technicians to obtain an overview of the status for each inclusion in terms of progress towards the sequencing end goal.

### 2.3. The PrOnco web platform

PrOnco was initially built to circumvent the limitations of REDCap in the context of the ProSeq project^9^ and was later extended to be able to support multiple precision oncology projects. It is developed around a core element containing a set of features common to all projects, and optionally, project-specific modules, if custom integration and tools are required. The core element provides utilities to leverage available external solutions through APIs, primarily REDCap, and includes a searchable variant database structured around patients, cases, and samples.

The modular approach limits the downtime of the whole system by decoupling each element to allow them to function independently (see Figure 4). This also facilitates the maintenance and upgrades of the system.

**Figure 4.**
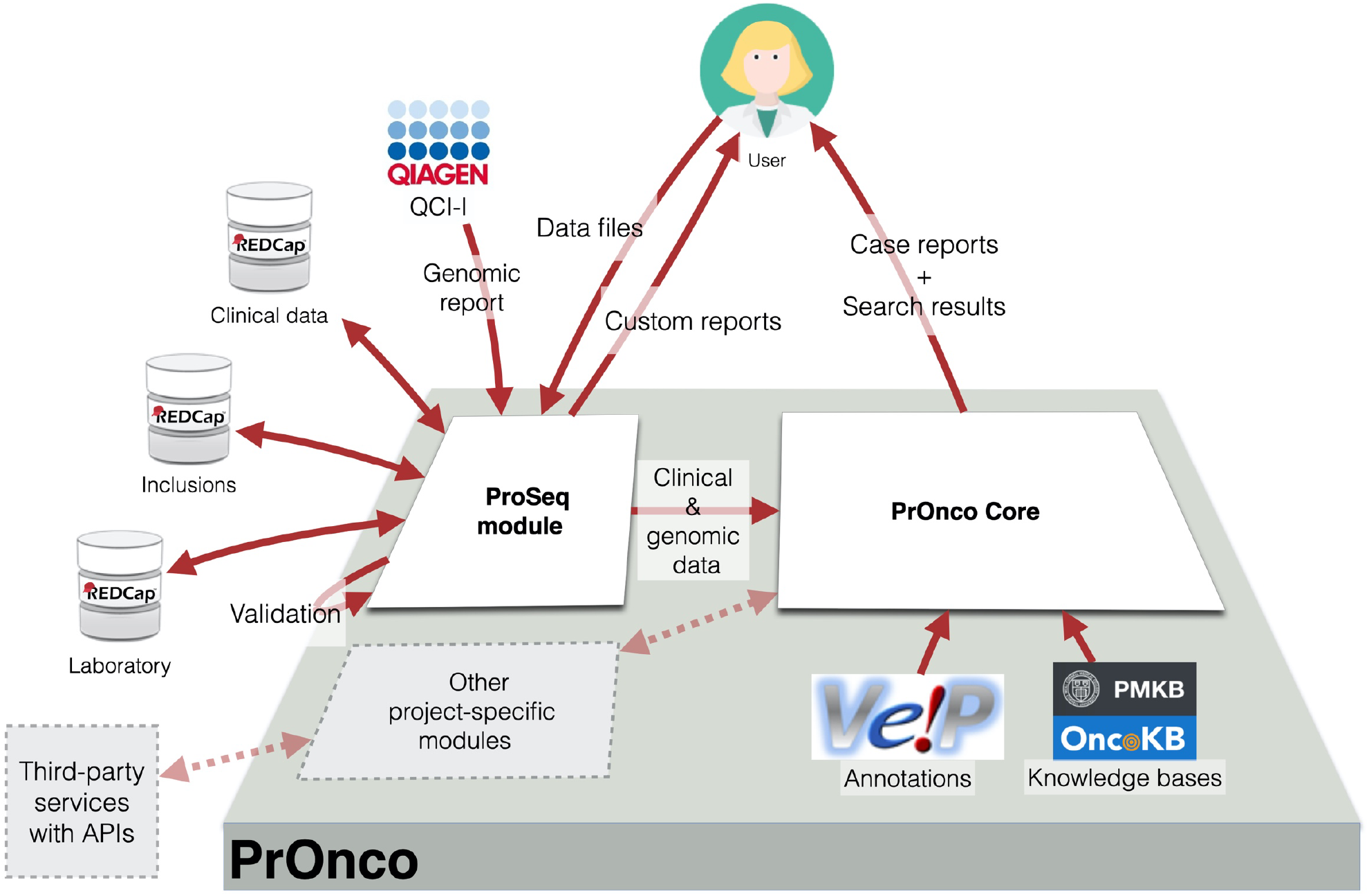
Overall PrOnco architecture. Arrows represent data flow. Except for user interactions (importing data files, accessing custom reports, and exploring the data), all flows of data are fully automated. The “Other project-specific modules” block illustrates the possibility of tightly integrating the platform with other projects. The Clinical data, Inclusions, and Lab REDCap databases are independent projects on a single REDCap server. For readability, only the ProSeq module has been represented. Variant Effect Predictor (VEP) software integrates other data sources, such as COSMIC17, dbSNP18, the 1000 Genomes Project19, or GnomAD20, which have not been represented here, again for readability. QCI-I is a commercial decision-support tool developed by QIAGEN.

PrOnco has built-in access control mechanisms that restrict access to data based on user profiles and projects as well as an audit trail to comply with European general data protection regulations (GDPR), since it displays patient-sensitive data.

PrOnco is developed using modern web technologies, notably the Python 3.8^21^ web framework Django 3.2^22^, Bootstrap 4^23^, and good practices in software development, including extensive unit testing and versioning through Git^24^. It also makes use of containerization with Docker^25^ to ease deployment and allow scalability.

The initial specifications for the platform were defined from needs expressed by clinical personal, molecular biologists, and bioinformaticians. The development process followed an agile methodology to allow the integration of early user feedback. It consisted of regular meetings with end-users to refine the roadmap and releases based on new or updated needs, translated into tasks in a Kanban board to be prioritized and assigned.

### 2.4. Data sources for genomic annotations

During the REDCap data synchronization, newly available VCF files are also imported to the platform. Since PrOnco is designed to support different projects, potentially using different reference genomes, the variant files can be lifted to the reference genome GRCh38^26^ using CrossMap software^27^. The purpose of this step is to simplify the downstream processing and homogenize the data sets.

To obtain a default set of annotations, each file is processed through the Ensembl Variant Effect Predictor (VEP) software^28^ (see Figure 3). VEP was integrated through a Docker container and a custom Python wrapper to communicate with the PrOnco core through a secure connection.

QCI-I was integrated with the platform through its API. Annotations performed on the QCI-I platform are retrieved automatically to generate the report directly on the PrOnco platform. Integration with QCI-I is, nevertheless, optional, and the classification of significance, reportability, and actionability can be added or edited manually on the platform. The actionability assessment follows international standards^29^.

Furthermore, annotations from OncoKB^30^ and PMKB^31^ are added dynamically when the list of variants is accessed to provide the user with additional background knowledge. These annotations, in addition to those provided by VEP, are available to all projects.

### 2.5. Tumour board report

The platform can provide, for each project, a comprehensive custom report to clinicians, incorporating both clinical and genomic data. In the context of the ProSeq module, we have developed a prototype for an interactive tumour board reporting interface (see Figure 5). It lists clinically relevant parameters (“Patient info” section), i.e., age, sex, body mass index, ECOG performance status, and comorbidities, to calculate Charlson’s comorbidity index, as well as the patient trajectory (“History” section) with each related cancer diagnosis, inclusion date, morphology, significant genomic alterations, and treatment regimen. Only limited clinical data are stored on the PrOnco platform, the rest are retrieved directly from REDCap.

**Figure 5.**
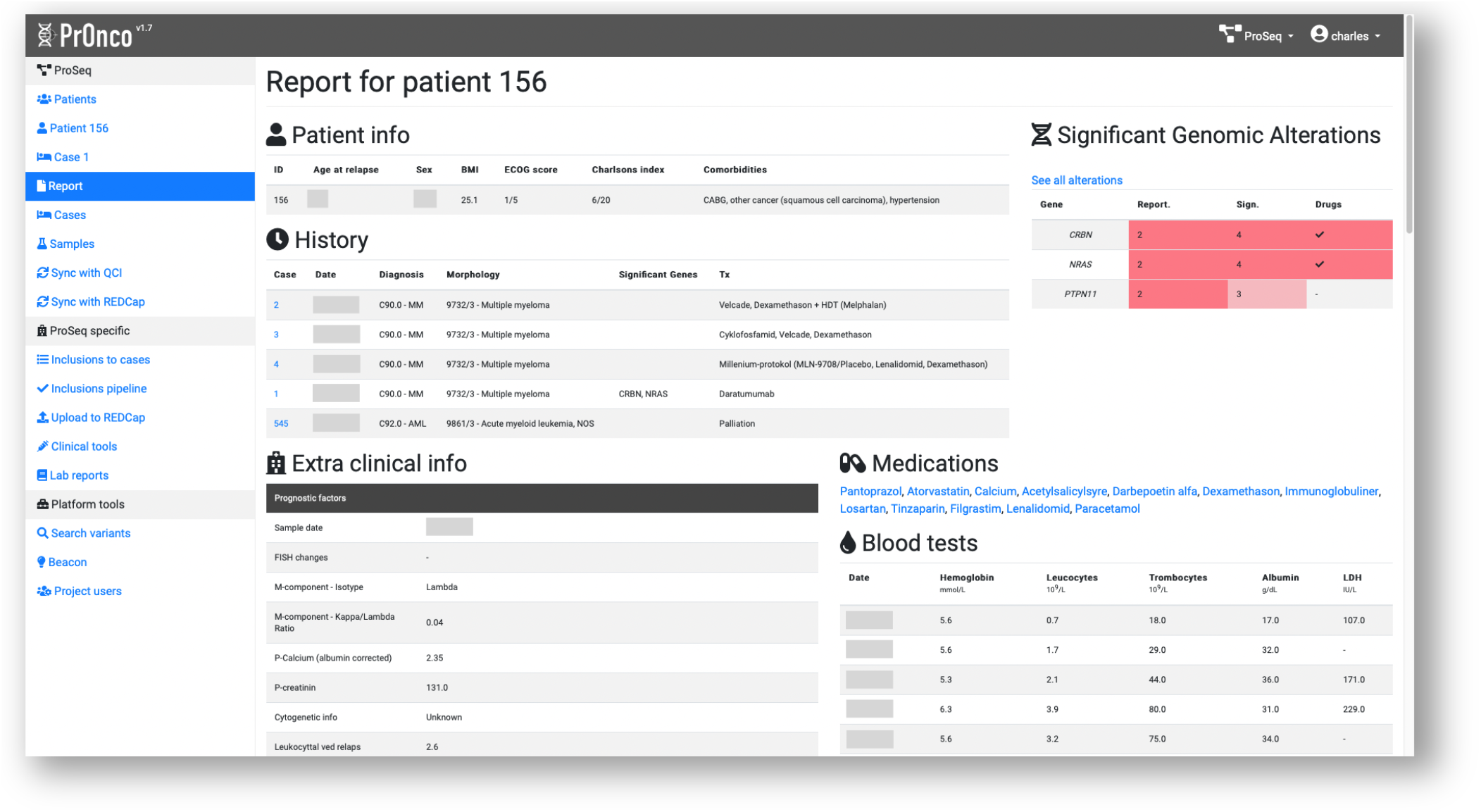
Prototype for an online tumour board report developed for haematological cancers. The interface can be customized based on available data. The description of the various elements in this screenshot can be found in the “Tumour board report” section.

A more detailed listing of the significant genomic alterations (“Significant genomic alterations” section) for the current case is also displayed based on a significance evaluation done either in QCI-I or by hand in the platform. From this list, detailed information can be obtained about each variant in overlay with the list of VEP transcripts and the significance, reportability, and actionability alongside notes and annotations from OncoKB and PMKB, if available.

These generated reports can be exported as a .PDF for printing or archiving.

The interactive reports have been presented internally to six haematologists, and their early feedback has been used to adjust the list and order of the presented clinical parameters.

### 2.6. Data findability

REDCap is intended as a data capture solution and has relatively limited search functionalities, notably for data stored as files. To improve the findability of the genomic data stored in REDCap, a user-friendly search interface has been integrated in PrOnco, which allows searching for variants based on genes, chromosomes, precise positions or ranges, reference alleles and alternative alleles, as well as clinical data such as age range, sex, and diagnosis. To further expand the findability of the genomic data at an international level, we have implemented the Beacon API to take part in the federated the Beacon Network^32^. The overall data workflow is presented in Figure 6.

**Figure 6.**
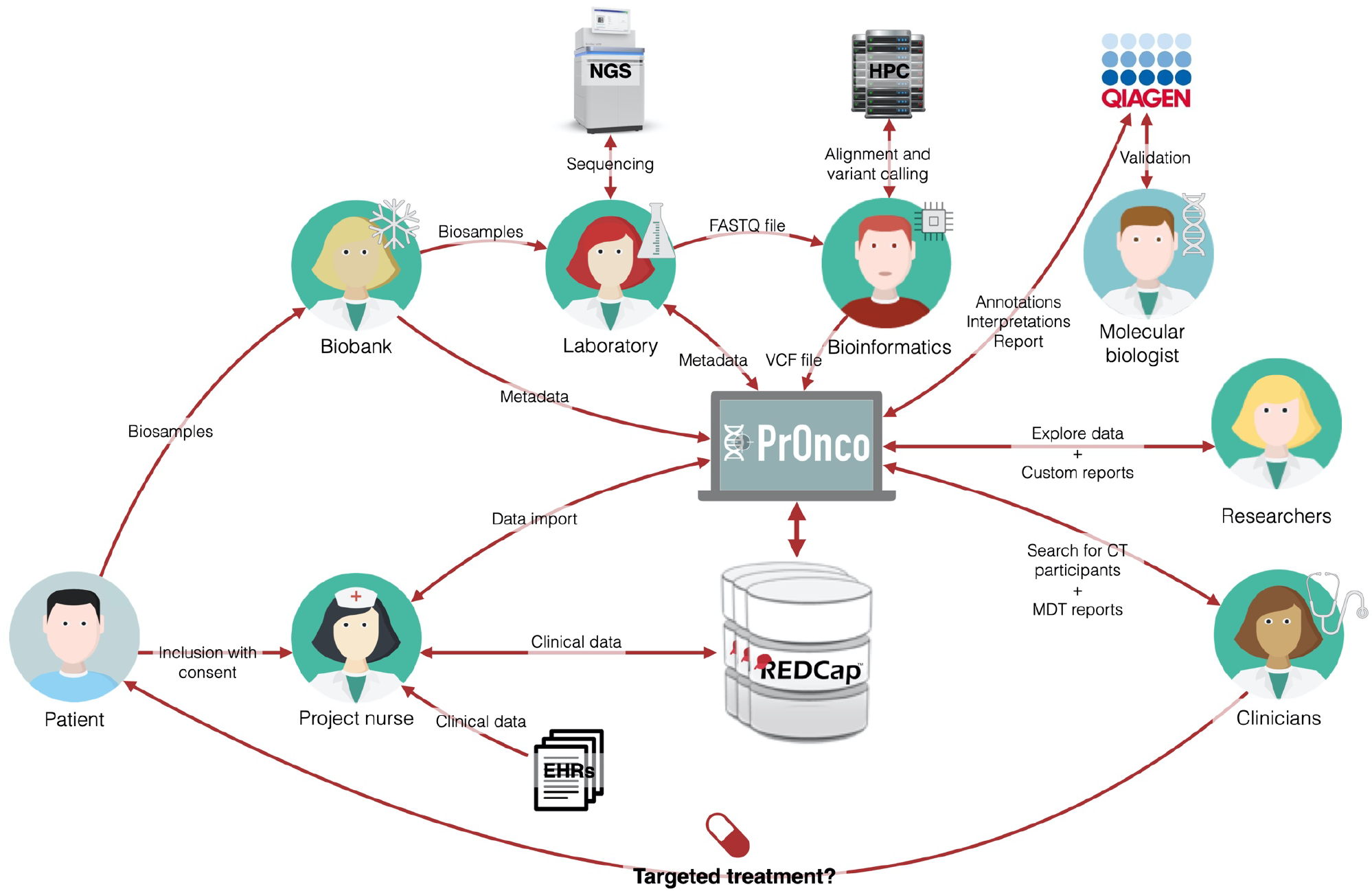
Overall data workflow for the ProSeq project. EHRs, Electronic Health Records; NGS, Next Generation Sequencing machine; HPC, High Performance Computing service.

## 3. USAGE

We present in the following an overview of data collected in REDCap across the four projects at the time of writing (December 31, 2021) and the usage of the platform, in relation to the three objectives initially defined. The results regarding the actionability of the genomic findings for the ProSeq projects have been presented in a previously published article^9^.

The usage was evaluated looking at the number of page views, the number of daily active users, and the number of monthly active users. The activity of the developers of the platform was ignored and only successfully loaded web page were considered. A monthly active user is a user that has connected at least once to the platform in a month. Similarly, a daily active user is a user that has connected at least once to the platform per day.

### 3.1. Collected data

For all four projects, data for a total of 1921 patients have been entered in REDCap covering a variety of diagnoses. (See Table 1).

**Table 1.** Distribution of clinical records per project and diagnosis.

| Diagnosis | Overall | ProSeq | RetroSeq | ProSeq Cancer | DCCC-Hem |
| --- | --- | --- | --- | --- | --- |
| MDS | 305 | 11 |  |  | 294 |
| AML | 258-266 | 30 | 1-5 | 1-5 | 226 |
| CCUS | 120 |  |  |  | 120 |
| DLBCL | 103 | 50 | 53 |  |  |
| CMML | 67-71 | 1-5 |  |  | 66 |
| MM | 49 | 49 |  |  |  |
| CLL | 44 | 44 |  |  |  |
| FL | 40-48 | 38 | 1-5 | 1-5 |  |
| Other haematological diagnoses | 796 | 94 |  | 6 | 696 |
| Total haematology | 1781-1801 | 316-320 | 55-63 | 8-16 | 1402 |
| Breast cancer | 27 |  |  | 27 |  |
| Lung cancer | 16 |  |  | 16 |  |
| Ovarian cancer | 15 |  |  | 15 |  |
| Prostate cancer | 15 |  |  | 15 |  |
| Brain cancer | 13 |  |  | 13 |  |
| Other oncological diagnoses | 48 |  |  | 48 |  |
| Total oncology | 134 | 0 | 0 | 134 | 0 |
| Total | 1915-1935 | 316-320 | 55-63 | 142-150 | 1402 |

*ICUS, idiopathic cytopenia of undetermined significance, MDS, myelodysplastic syndrome, AML, acute myeloid leukaemia, DLBCL, diffuse large B-cell lymphoma, CMML, Chronic myelomonocytic leukaemia, MM, multiple myeloma, CLL, Chronic lymphocytic leukaemia, FL, follicular lymphoma. Ranges are used to preserve anonymity where individual values are below 6.*

Data from 1,980 patients were stored in PrOnco, corresponding to 447 genomic data files. From these data, 489,695 unique variants have been registered.

These data have been used internally to evaluate readily available cohort sizes for research, focusing on specific sets of genes. The clinical data collected were also used in projects dealing with health economy and biological mechanisms.

### 3.2. Platform usage

From February 2019 to December 2021, the four active projects had 36 active users on the platform and, on average, 7.9 monthly active users and 1.8 daily active users.

In terms of page views, PrOnco was primarily used to explore the available data and was secondarily used to obtain overviews of the data and workflows (see Table 2). The features developed to facilitate the import of data and control its quality represented more than 25% of the views.

**Table 2.** Page views per category. Page views for accessing the platform (login page, home page, etc.) are not included here.

| Category | Page views (% of total) | Description |
| --- | --- | --- |
| Data exploration | 1,588 (34%) | Listing and information about specific projects, patients, cases, samples, and files |
| Workflow reporting | 1,162 (28%) | Reporting views for inclusions, laboratory work, and clinical data collection |
| Data import | 621 (16%) | Import and migration of biobank, pharmacy, and transfusion data |
| Data validation | 398 (10%) | Synchronization and validation of REDCap data |
| Tumour board reports | 133 (3%) | Interactive report generation for tumour board meetings |
| Data search | 104 (3%) | Search interface based on genomic and clinical data |

The data import and validation features were nevertheless extensively used, reducing the manual workload for the capture of these data, and potentially improving the quality of the collected data by identifying inconsistencies.

## 4. DISCUSSION

### 4.1. Advantages and limitations of REDCap

REDCap is a popular solution for data capture in health science and has been widely adopted across the globe and especially in Denmark where virtually all university hospitals host a REDCap server. Notably, the audit-trail mechanism makes it GDPR compliant and thus more easily approved for storing potentially sensitive patient information. Creating a data repository is relatively straight-forward for a standard clinical trial, with design tools making it accessible to people with limited IT skills. REDCap’s built-in control mechanisms allow admins to precisely define what users can see and export. This greatly improve the accessibility of data, allowing for a controlled web access to the data and thus helps making the data FAIR. But the current version of REDCap cannot be used as an end-to-end data solution due to its limitations in terms of support of complex data structure, reporting, and integration with third party solutions, requiring specific developments.

To be searchable, data are typically stored in a relational database where entities are connected through foreign keys. It is therefore possible to build complex relational structures for the data. Such principles can be reproduced using REDCap as a data storage but only in a very limited way using the instance concept. Building more complex data structure requires to create separated REDCap repositories, as illustrated in Figure 1 for the ProSeq project, but with the drawback of having to handle lose relations between entities, increasing the risk of errors without external validation.

The genomic data is generated as VCF files which can contain large amounts of data, that are difficult to store directly in REDCap, especially if annotations are added. It was therefore decided to store these data directly as flat files. This solved the problem of data complexity, but it also prevented these data from being findable without additional developments.

Concerning the reporting, while some features are available on REDCap through custom reports, presentation options are limited. Furthermore, combining data from multiple repositories, needed in the case of complex projects such as the ProSeq project requires additional developments.

Finally, integrating third party components cannot be done in a simple manner directly in REDCap and therefore requires specific developments to manage the connections between components.

These three limitations can be alleviated thanks to an external platform to allow for a proper end-to-end solution.

### 4.2. The PrOnco platform

PrOnco is designed as a modular structure to reuse, wherever relevant, existing solutions that fulfil a specific need. Furthermore, the decoupling of elements of the infrastructure allows them to work independently to limit overall downtime, e.g., REDCap will stay available while PrOnco is being updated. Building a larger scale platform, encompassing the data capture parts, gives a lot of flexibility in terms of features and data structure but potentially lose the decoupling between the data capture and data usage and introduces a high upfront cost to replicate existing features in REDCap. Using a smaller platform in addition to REDCap also allows for a quick start with data collection, using a reliable data capture solution, while the data usage part can be built incrementally.

Another solution would have been to use a wider-scope existing solution such as the one developed by Kang et al.^6^, but can be challenging to integrate and customize and miss some of the features available in REDCap.

A customized solution always raises the question of long-term maintenance, a problem that is prevalent in academic software development. Our goal was to make the platform sufficiently project-agnostic to be used in a variety of projects over the years, which would allow long-term support.

A questionable choice was to develop a variant database on the platform. Indeed, solutions such as cBioPortal^33^ could have been used to make the genomic data searchable on the platform. We nevertheless decided to build our own solution to be able to handle specific annotations, notably from VEP and QCI.

More generally when developing a custom platform, the question is when to use existing solutions or develop new ones. This is notably the case with the LIMS backbone which was developed using REDCap and custom features on the platform for the ProSeq project while a commercial solution, like Titian’s Mosaic system^34^, was used for the DCCC-Haem project. At the end, it boils down to how well existing solutions fit the need, how expensive they are, and what is already in place.

### 4.3. The FAIR data principles

The accessibility can be handled directly into REDCap by giving access to specific parts of the repositories to researchers. The interoperability and reusability can be achieved through the REDCap data structure, with the encoding of values using international standards and the presence of sufficient metadata.

However, the findability is not addressed on REDCap, because it is extremely specific to the context of the project, the data might not be stored in a format easily searchable, and a typical limiting factor to find these types of data is a lack of bioinformatics know-how. By making it available through a user-friendly interface, we allow, for example, direct investigation of the feasibility of a study according to available data for the researchers.

Thanks to the built-in search functionality and the Beacon interface, we made these data findable to facilitate research and participate in global data-sharing initiatives.

### 4.4. Perspective

Through use of the platform, needs have emerged that will have to be addressed. For example, currently it only supports genomic data from VCF files, i.e., single-nucleotide variants and small insertions and deletions. This can be a problem in neoplastic disorders, which are often related to larger structural variants, but we plan to support these types of data in the future.

Another key aspect for data collection, which is not yet incorporated in the platform, is the capability of retrieving data automatically from EHRs which currently relies heavily on manual work. Import tools are a step towards the automation of clinical data retrieval, to streamline the workflow, reduce human errors and workload, and thereby costs. But depending on the current EHR systems in place, this will require, potentially extensive, but necessary IT work. Contact has been taken with EHR solution providers to explore the technical and financial feasibility of these developments.

For the laboratory part, the solution could potentially be expanded to support advanced integration with barcode-based sample processing traceability.

Finally, improvements should be planned to build a tumour board report as relevant as possible for clinicians. The proof-of-concept prototype for an interactive tumour board report has been presented in a close-to-clinical context for a sample of currently treated patient cases by haematologists. This tumour board report is still in the prototyping phase, but it allows us to explore the feasibility of such a concept in a Danish context and to identify bottlenecks and roadblocks to make it usable in a clinical context. The report must contain genetic data, summarized in a clinically meaningful manner, that are assessed, interpreted, and prioritized by experienced personnel to support clinical treatment guidance. Furthermore, an automatic search function pairing individuals with criteria for treatment in protocols on a national level should be developed.

Another aspect to take into consideration is the handling of the reclassification of variants. Indeed, knowledge about the impact of variants is growing continuously and variants of unknown significance might be reclassified. In a clinical context, these changes should be tracked for quality purposes.

## 5. CONCLUSION

We have developed an end-to-end modular precision oncology solution, from clinical data collection to the generation of tumour board reports, leveraging existing solutions, notably REDCap. With limited resources, this allowed us to improve the quality and efficiency of the collection of both clinical data and genomic data and to make the data generated more FAIR. The collected genomic data were made available internally and externally to support cancer research. The prototype of an interactive tumour board report allowed us to explore the feasibility of its implementation in a Danish clinical context. More work is, nevertheless, required for full integration with electronic health records, support for more types of genomic data, and to validate the tumour board report for both haematology and oncology. However, for both clinicians and researchers, being able to combine a large amount of clinical and genomic data and make it easily accessible is of great value for their work, and continuous effort should be made to make this a reality.

## Data Availability

The data mentioned in this present work were collected in the context of other research projects, their accessibility is therefore defined by these projects.

## AVAILABILITY

The REDCap data dictionaries for all repositories for all projects can be made available upon request. For the platform, a free user license can be granted for non-profit and public organizations. If you are interested in collaboration on PrOnco development, please contact the main author. Data for all projects are findable on the Beacon Network central node, here is a search example: https://beacon-network.org/#/search?pos=7674220&chrom=17&allele=T&ref=C&rs=GRCh38

## AUTHORS’S CONTRIBUTIONS

All authors have made a substantial, direct, intellectual contribution to this study.

## ACKNOWLEDGEMENTS

We would like to thank the following for their contribution and feedback: project nurse Pernille From Blindum; biobank and laboratory technicians Jane Ildal, Anja Führer, Helle Høholt, and Louise Hvilshøj Madsen; software developers Anders Sune Pedersen, Dennis de Weerdt and Louise Pedersen Pilgaard; MDs Henrik Gregersen, Jakob Madsen, Peter D. Jensen and Paw Jensen from the Department of Haematology, Aalborg University Hospital; molecular pathologist Lykke Grubach from the Department of Pathology, Aalborg University Hospital; the North Denmark Region IT department; the members of the NEXT Bioinformatics Group and the members of the Danish Clinical Bioinformatics Group (DCBIG).

